# Interpretable Automated Detection and Measurement of Dome-Shaped Macula on Optical Coherence Tomography

**DOI:** 10.64898/2026.09.28.26356190

**Authors:** Fan Yang, Yanping Zhou, Bo Zhang, Jinliuxing Yang, Linlin DuMPH, Chunxia YuMMed, Xia Chen, Jianfeng Zhu, Yulan Wang, Xiangui He

**Author notes:** Co-corresponding authors: Yulan Wang and Xiangui He. Commercial Relationships Disclosure:* F. Yang, None; Y. Zhou, None; B. Zhang, None; J. Yang, None; L. Du, None; C. Yu, None; X. Chen, None; J. Zhu, None; Y. Wang, None; X. He, None.

## Abstract

**Purpose:** To develop an interpretable automated pipeline for dome-shaped macula (DSM) detection and measurement from optical coherence tomography (OCT) B-scans.

**Methods:** We analyzed one eye from each of 300 children aged 4–18 years in the Shanghai Child and Adolescent Large-scale Eye Study High Myopia (SCALE-HM) cohort (50 DSM-positive; 100 test eyes). A U-Net pretrained on OCTA-500 and fine-tuned on the development set segmented the retinal pigment epithelium (RPE) boundary. A geometric module located the apex, fitted a tangent-line baseline, and computed perpendicular height in physical coordinates.

An eye was DSM-positive if any gradable B-scan had height *≥* 50 µm. Two masked graders evaluated the test set; a third adjudicated disagreements.

**Results:** The area under the receiver operating characteristic curve was 0.927 (95% confidence interval [CI], 0.850–0.986), sensitivity was 82.1% (95% CI, 64.4%–92.1%), and specificity was 100% (95% CI, 94.9%–100%) against the adjudicated reference. In concordant-positive eyes, automated height agreed with manual measurements (intraclass correlation coefficient, 0.949; mean absolute error, 6.9 µm; bias, *−*5.7 µm, versus Doctor A). Mean RPE-boundary error on pediatric test scans was 1.54 µm.

**Conclusions:** The pipeline may be considered a reliable and reproducible tool for automated DSM detection and height measurement on radial OCT, with each value traceable to landmarks on the source B-scan. External validation across populations, devices, and acquisition protocols is needed.

**Translational Relevance:** The pipeline provides an objective and reproducible method to detect DSM and quantify dome height, width, and orientation across OCT meridians. Because each measurement can be verified on the source B-scan, it enables standardized DSM assessment.

---

Dome-shaped macula (DSM), first described by Gaucher et al. in 2008,^1^ was originally characterized as an inward macular convexity within a posterior staphyloma in highly myopic eyes. Current OCT-based definitions are broader and encompass a dome-like anterior protrusion of the retinal pigment epithelium (RPE) line that may occur without posterior staphyloma and across a wider refractive spectrum.^2–5^ DSM is clinically relevant mainly because of its associated complications, particularly serous macular detachment and RPE atrophy, which can threaten central vision, whereas its relationships with choroidal neovascularization, choroidal thickness, and visual function are more heterogeneous.^5–8^ Recent radial-scan studies have proposed distinguishing complete round DSM from ridge-shaped macula (RSM), in which the elevation is confined to selected meridians, although this terminology is not yet uniformly applied.^5,9^ Because DSM is defined by the shape of a single OCT boundary, its detection and grading depend directly on how that boundary is measured.

In clinical practice, DSM is identified by subjective visual inspection of the OCT B-scan, and dome height, when it is recorded at all, is measured manually on the scan. This workflow has three main limitations. First, detection depends on each ophthalmologist’s impression of whether the RPE curves inward relative to the surrounding posterior pole, which produces substantial intergrader variability. This problem is greatest for shallow domes close to the diagnostic threshold, where small differences in apex or baseline placement can change the classification. Second, although manual height measurement does yield a numeric value, its reproducibility and standardization remain limited. Within a study, apex and baseline placement on noisy OCT images depend on the operator. Across studies, baseline definition and placement vary, with reported approaches ranging from a line joining the lowest points of the staphyloma to a chord between the parafoveal inflection points, so prevalence estimates and measurements can be difficult to compare between reports.^5,6^ Third, assessment is usually done on a single B-scan orientation, typically horizontal or vertical, which can miss asymmetric domes or underestimate the maximum dome height when the peak lies along another meridian. Even with a multi-orientation radial protocol, inspecting every meridian by hand is slow and still subjective, making comprehensive manual assessment impractical for longitudinal monitoring of growing eyes or for large-scale studies.

Automated approaches to DSM assessment have begun to emerge but remain limited in scope. Del Fabbro et al.^10^ proposed the dome-shaped macula curvature (DSMC) index, which combines inflection-point chord length with dome height and showed high inter-grader reliability. Their workflow evaluates the available scans and selects the scan with the highest curvature, but still requires manual landmark identification in proprietary software and models the dome with idealized shapes rather than tracing the full RPE contour. Ye et al.^11^ demonstrated the feasibility of deep learning for DSM detection using an end-to-end classifier in pathologic myopia, but their model inherits the subjectivity of its training labels and outputs only a binary decision, with no continuous, verifiable measurement. The attention maps they provide highlight salient image regions but do not recover the landmarks from which dome height is defined. Interpretable, segmentation-based OCT pipelines have nonetheless proven feasible in other retinal diseases.^12–14^ What remains missing is an automated method that measures DSM reproducibly, ties each measurement to identifiable structures on the source scan, and systematically summarizes all meridians of a radial acquisition. We therefore developed a deterministic, anatomy-traceable pipeline and evaluated it in a pediatric high-myopia cohort as a controlled clinical imaging test bed.

Our primary aim was to develop and evaluate an interpretable automated pipeline for DSM detection and measurement from OCT B-scans. The pipeline measures each B-scan independently and summarizes all available meridians; in this study, it was evaluated on 12-line radial acquisitions from a pediatric high-myopia cohort, used as a controlled clinical imaging test bed, with masked expert grading on a held-out test set. The pipeline was designed so that each numeric output is derived from identifiable structures on the source B-scan, allowing direct clinical verification and consistent comparison across scans.

## Methods

### Study Design and Participants

This retrospective study adhered to the tenets of the Declaration of Helsinki and was approved by the ethics committee of the Shanghai Eye Disease Prevention and Treatment Center. Participants were drawn from the high myopia registration study of the Shanghai Child and Adolescent Largescale Eye Study (SCALE-HM), a prospective, examiner-masked study. As described in our previous methodological articles, SCALE-HM participants were selected from the SCALE study,^15^ a city-wide, school-based study covering over one million children in Shanghai, and the detailed SCALE-HM protocol has been published.^16^ Before the original study began, its rationale was explained to all children and their parents or guardians. Written informed consent was obtained from all parents or guardians and from children aged 12 years or older, and oral consent was obtained from children younger than 12 years. Because this was a retrospective image-analysis study, the institutional review board waived the requirement for additional consent. Participants self-identified, or were identified by their parents or guardians for younger children, as Han Chinese.

Children and adolescents aged 4 to 18 years were eligible if they met the age-specific definition of high myopia and had a gradable fovea-centered radial OCT. The cutoff for high myopia was set separately for each age band, reflecting the fact that refractive error in childhood drifts from hyperopia toward myopia as the eye matures, so that a degree of myopia that is unremarkable in a teenager is abnormal in a young child. Using spherical equivalent (SE, the spherical power plus half the cylindrical power), the thresholds were SE *≤ −*0.5 D for ages 4 to 5, SE *≤ −*3.0 D for ages 6 to 8, SE *≤ −*5.0 D for ages 9 to 12, and SE *≤ −*6.0 D for ages 13 and above.^16^ Refraction was obtained under cycloplegia following the SCALE-HM protocol.^16^ Eyes were excluded for organic ocular disease (strabismus, moderate to severe ptosis, congenital cataract, or glaucoma), any fundus disease other than myopia-related lesions, previous intraocular or refractive surgery, or OCT image quality too poor for reliable delineation of the RPE.

The cohort was assembled by design rather than by random population sampling, to span the full range of DSM severity. DSM-positive eyes were first identified during clinical diagnostic screening and were then manually re-graded and re-measured by a single experienced grader, yielding 50 DSM-positive eyes with dome heights from 50 to 248 µm, together with 250 DSM-negative eyes, giving a cohort of 300 eyes from 300 patients. One eye per patient was included to avoid within-subject correlation, with random selection when both eyes were eligible. For consistency with prior OCT studies, an eye was considered DSM-positive when the RPE outer border showed an inward convexity with a dome height *≥* 50 µm on one or more B-scans.^5,17^ Throughout this study, DSM is used as an umbrella OCT term; configurations in which the threshold-crossing elevation was confined to selected meridians, described by some authors as RSM, were retained and characterized by orientation rather than treated as a separate diagnosis.^9^ The 300 eyes were split into a 200-eye development set, used for segmentation model training (160 eyes) and internal validation (40 eyes), and a 100-eye test set drawn at random. The DSM status of the test set was re-determined independently by the three-grader protocol described below, which served as the reference standard for evaluating the pipeline. The single-grader cohort labels were used only for the exploratory morphological analysis. Figure 1A illustrates the cohort construction and split.

**Figure 1:**
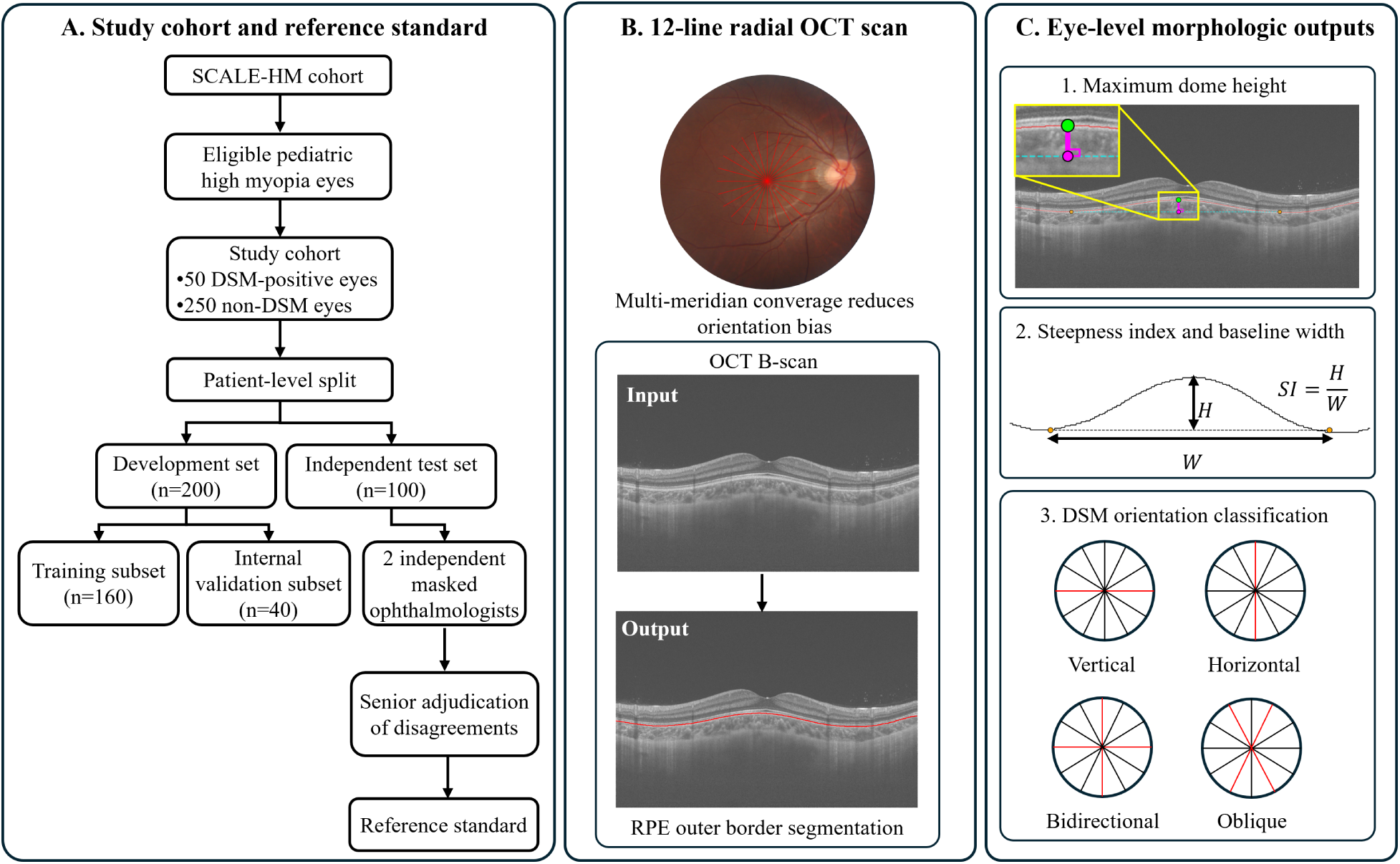
Study design and automated DSM analysis pipeline. **A,** Cohort construction and patient-level split: 300 eyes (50 DSM-positive, 250 DSM-negative) were drawn from the SCALE-HM cohort and split into a 200-eye development set (160 training, 40 internal validation) and a 100-eye independent test set. The test set was reviewed independently by two masked graders, with disagreements resolved by adjudication. **B,** 12-line radial OCT acquisition centered on the fovea provided multi-meridian input to the segmentation-then-geometry pipeline. **C,** Eye-level morphologic outputs derived across all meridians: maximum dome height (*H*), maximum projected base width (*W*), steepness index (*SI* = *H/W*), and DSM orientation classification. DSM = dome-shaped macula.

### Image Acquisition and Reference Standards

OCT imaging was performed using a swept-source OCT device (DRI OCT Triton, Topcon Corporation, Tokyo, Japan). A 12-line radial scan protocol was used, with B-scans acquired through the fovea at equally spaced meridians (0*^◦^*, 15*^◦^*, 30*^◦^*,…,165*^◦^*). This acquisition scheme (Figure 1B) enables identification of the maximum dome height across orientations and characterization of dome orientation, and serves as the input to both manual review and automated analysis. Baseline clinical variables, including age, axial length, spherical equivalent, best-corrected visual acuity, intraocular pressure, and anterior chamber depth, were extracted from the electronic medical records to describe the evaluated cohort. Lateral and axial sampling intervals (8.789 and 2.609 µm per pixel, respectively) were read from the scan metadata and used for physical-coordinate conversion. All manual annotations followed a three-grader protocol involving three attending ophthalmologists with 10 to 16 years of clinical experience. For each annotation task, two of them graded every eye independently and the third adjudicated any disagreement. Two annotation tasks were performed: one provided the eye-level and scan-level reference standard used to evaluate the complete pipeline, and the other provided the pixel-level RPE ground truth used to train and test the segmentation model.

### DSM and dome height grading

On the 100-eye test set, two of the ophthalmologists independently reviewed all 12 B-scans per eye, masked to the algorithm and to each other. Each first made a binary DSM judgment, then for DSM-positive eyes selected the B-scan(s) with the most prominent dome and measured its height with a custom ruler tool. To do so, the grader placed a straight baseline touching the basal RPE on both sides of the dome and marked the apex, and the tool recorded the perpendicular distance from the apex to that baseline in micrometers. This baseline construction mirrors the tangent-line baseline used by the automated pipeline, supporting direct comparison between manual and automated measurements. Manual heights were used as recorded, without post hoc recalibration to the automated physical-coordinate definition. When the two graders disagreed on DSM status or on the selected dome height, the third ophthalmologist reviewed the B-scans and adjudicated the final label. The adjudicated assessments served as the reference standard for both eye-level DSM detection and scan-level dome height measurement on the test set. Algorithm performance was also reported against each grader individually as a sensitivity analysis.

### RPE outer border annotation

For segmentation model training and evaluation, the RPE outer border was manually annotated on one B-scan in each of 250 eyes, comprising all 200 development-set eyes and 50 of the 100 test-set eyes, yielding 160 training, 40 validation, and 50 test annotations. For DSM-positive eyes this was the dome-containing B-scan, and for DSM-negative eyes the horizontal B-scan. Two of the ophthalmologists independently reviewed and corrected an automated initial segmentation produced by OCTSEG,^18^ an open-source MATLAB tool for retinal layer segmentation that is independent of the deep learning pipeline used in this study. The third reviewed regions where the two annotations disagreed and set the final boundary positions. The mean absolute boundary error (MAE) between the algorithm and this consensus reference served as the segmentation accuracy metric on the 50-scan pediatric test subset.

### Automated Analysis Pipeline

The automated pipeline comprised two sequential components: segmentation of the RPE outer border, followed by geometric quantification of DSM morphology (Figure 1C).

### RPE Outer Border Segmentation

Accurate delineation of the RPE outer border was essential because all downstream DSM measurements were derived from this single boundary. Highly myopic pediatric eyes differ anatomically from those in public OCT datasets and may show pronounced posterior-pole curvature, retinal thinning, and, in some eyes, posterior staphyloma. We therefore adopted a two-stage transfer learning strategy. We used a standard U-Net^19,20^ without an ImageNet-pretrained encoder backbone, consisting of a contracting encoder with an initial convolutional block and four downsampling stages (64 to 1,024 feature channels) and a symmetric expanding decoder with transposed-convolution upsampling, linked by skip connections; each block consisted of two 3 *×* 3 convolutions with batch normalization and ReLU activation. It was first pre-trained on the public OCTA-500 dataset^21^ (OCTA-3M subset) to learn general retinal layer anatomy, then fine-tuned on the 160-eye pediatric training subset, with the 40-eye internal validation subset used for hyperparameter selection and early stopping. OCTA-500 splits were constructed at the volume level to prevent leakage between adjacent B-scans. Segmentation accuracy was reported out-of-sample, on both the OCTA-500 held-out split and the held-out pediatric test scans. Input B-scans were converted to grayscale, resized to 640 *×* 400 pixels (height *×* width), and scaled to [0, 1] by division by 255. A final 1 *×* 1 convolution produced the retinal layer segmentation, from which only the RPE outer border was used in this study; it was extracted as a one-pixel-wide boundary curve by a fixed post-processing step.

### Model Training and Hardware

Both the pre-training and fine-tuning stages were trained with a combined cross-entropy and Dice loss (equal weights) optimized by Adam (initial learning rate 1 *×* 10*^−^*^4^, weight decay 1 *×* 10*^−^*^5^, batch size 8). The learning rate was reduced by half whenever validation loss failed to improve for 5 consecutive epochs, and training was terminated by early stopping after 15 consecutive non-improving epochs (maximum 100 epochs). No data augmentation was applied. The model was implemented in PyTorch (version 2.10.0) and run on a single NVIDIA GeForce RTX 4090 GPU, with segmentation inference taking approximately 8 ms per B-scan.

### Geometric Quantification of DSM Morphology

On each radial B-scan, the segmented RPE outer border was smoothed with a Savitzky-Golay filter^22^. Candidate apex and tangent-contact points were selected from the smoothed contour using fixed deterministic rules, and the resulting pixel coordinates were mapped to physical coordinates using the lateral and axial sampling intervals recorded for the scan. If *p* = (*x, y*) denotes an image point, the physical point was (*s_x_x, s_z_y*), where *s_x_* and *s_z_* are the lateral and axial sampling intervals. Dome height was then calculated as the signed Euclidean perpendicular distance from the physical apex to the physical tangent-line baseline; the perpendicular foot was obtained by orthogonal projection onto the same baseline. Scans without a clear convex protrusion at the apex were excluded. Because the lateral and axial sampling intervals differ, computing the distance in physical coordinates avoids the error that arises when a sloped baseline is measured in pixel units and only afterward scaled by the axial spacing.^23^ It does not require explicit localization of the lowest staphyloma points or parafoveal inflection points and remains stable when these landmarks are ill-defined or asymmetric. Using the commonly applied 50 µm cutoff, an eye was classified as DSM-positive if at least one B-scan reached this height.^5,17^

Because each per-scan measurement is automated, it could be applied to all 12 meridians and reduced to a compact eye-level description of the dome. For each DSM-positive eye, three continuous features were computed from the per-meridian measurements: maximum dome height (the largest perpendicular apex-to-baseline distance across meridians), maximum projected base width (the largest horizontal extent between the two baseline contact points across meridians), and steepness index (maximum dome height divided by maximum projected base width).

The 12-meridian radial acquisition let us classify dome orientation across the full range of axes, including the oblique ridges that conventional horizontal-and-vertical scanning cannot resolve. Because a ridge appears as a dome only when a B-scan transects it perpendicular to its axis, orientation was inferred from the meridians on which a dome (*≥*50 µm) was detected. Defining near-horizontal B-scans as 0*^◦^ ±* 15*^◦^*and near-vertical as 90*^◦^ ±* 15*^◦^*, each DSM-positive eye was classified as vertical (dome only on near-horizontal scans, i.e., a vertically oriented ridge), horizontal (dome only on near-vertical scans, i.e., a horizontally oriented ridge), bidirectional (dome on both near-vertical and near-horizontal scans, corresponding to the round type of Caillaux et al.), or oblique (dome only on oblique scans, indicating a diagonal ridge). These labels describe meridional detection patterns; bidirectional positivity was not assumed to establish a complete round dome in all 12 meridians. This scheme adapts prior morphological categories to the full 12-meridian coverage.^6,9^ Representative B-scans for each pattern are shown in Figure 2.

**Figure 2:**
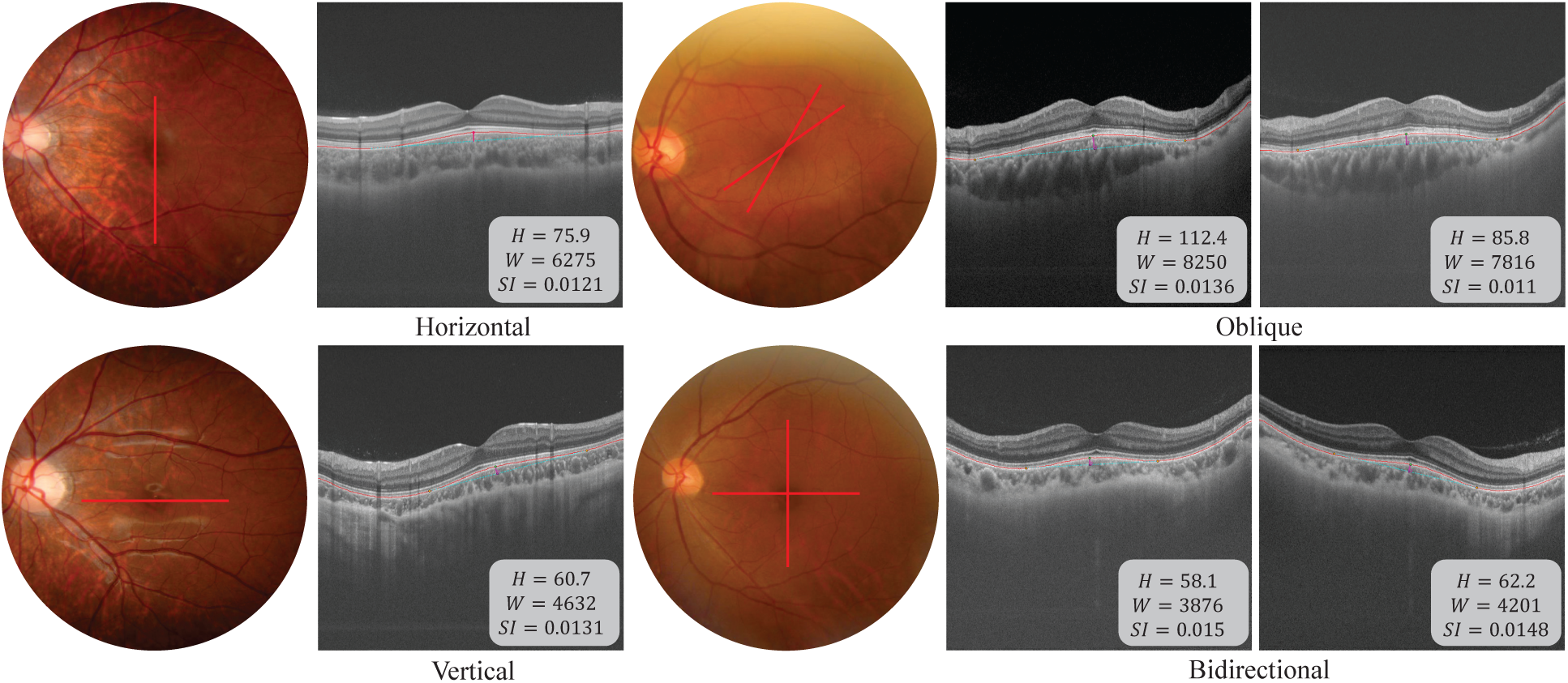
Representative DSM orientation patterns identified by the automated pipeline. Each case shows a fundus photograph with the dome-positive meridian(s) overlaid in red, together with the corresponding OCT B-scan(s) and per-scan measurements: maximum dome height (*H*, µm), maximum projected base width (*W*, µm), and steepness index (*SI* = *H/W*). **A,** Horizontal-oriented DSM (dome on the near-vertical meridian only). **B,** Oblique DSM (dome on two oblique meridians). **C,** Vertical-oriented DSM (dome on the near-horizontal meridian only). **D,** Bidirectional DSM (dome on both near-vertical and near-horizontal meridians).

### Interpretability Output

To support interpretability, the pipeline generated visual outputs at both the segmentation and geometric-analysis stages. The intermediate output displayed the retinal layer segmentation and derived RPE outer border overlaid on the original OCT B-scan. For dome-positive scans, the final analysis report additionally overlaid the geometric baseline, basal support points, apex, and perpendicular height line on the original image, enabling direct clinician verification of how each automated DSM measurement was derived from the source anatomy. Figure 3 illustrates the full pipeline on a representative B-scan.

**Figure 3:**
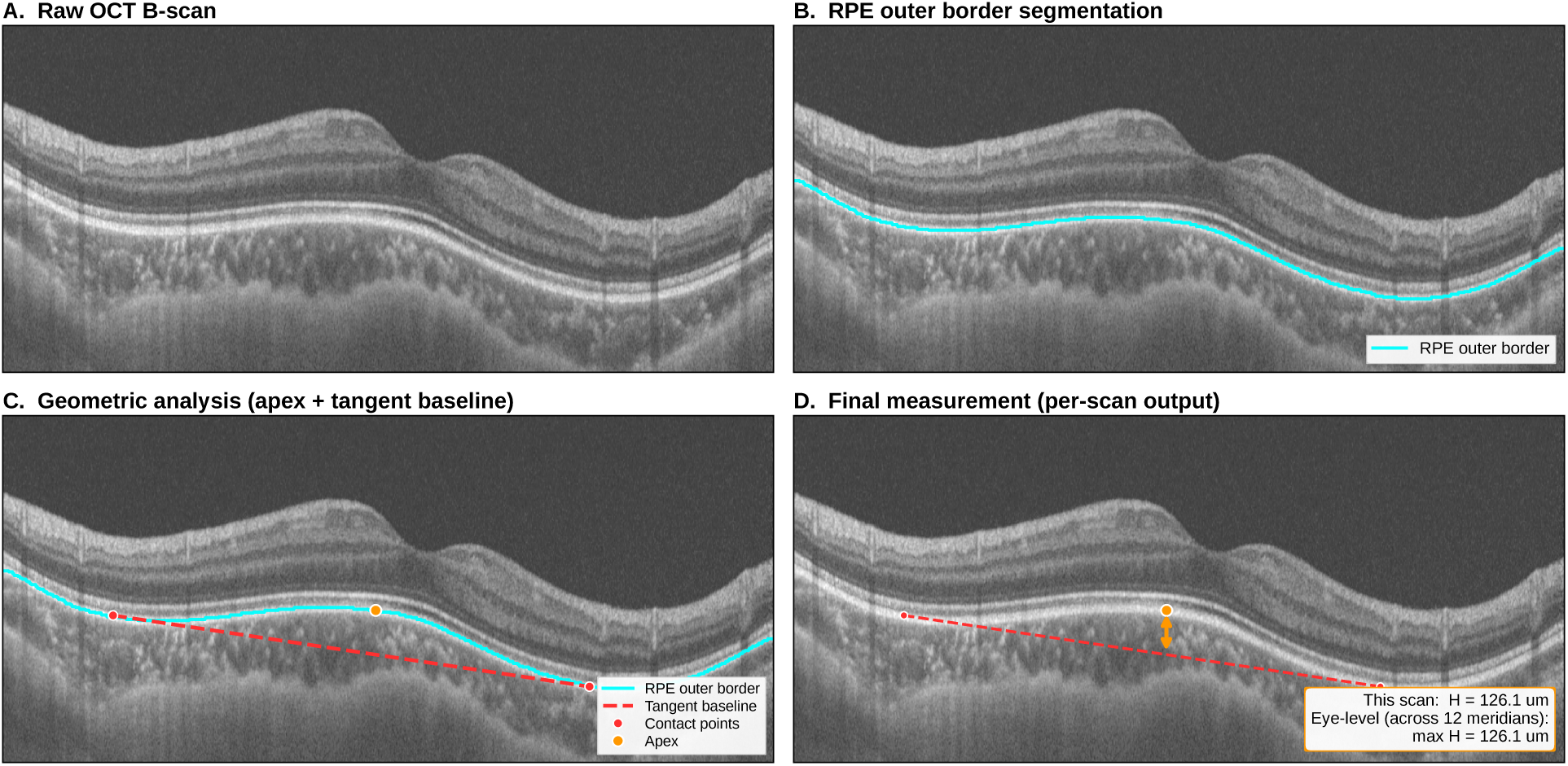
Step-by-step illustration of the automated DSM analysis pipeline on a representative B-scan. **A,** Raw OCT B-scan input. **B,** U-Net segmentation of the RPE outer border (cyan). **C,** Geometric analysis: dome apex localized on the smoothed RPE curve, and a tangent-line baseline (red dashed) fitted to the two contact points flanking the apex (Methods, §2.3). **D,** Final measurement: perpendicular distance from apex to tangent baseline (orange) is recorded as the scan’s dome height.

## Statistical Analysis

Continuous variables were summarized as mean *±* standard deviation and categorical variables as count (percentage). Baseline characteristics were compared using the Mann-Whitney *U* test for continuous variables and the chi-square test for categorical variables.

Three categories of outcomes were pre-specified: a primary outcome (eye-level DSM detection), a secondary outcome (scan-level dome height agreement), and supporting outcomes (segmentation accuracy and descriptive morphology). The analytic approach for each is detailed below.

For the primary outcome of eye-level DSM detection, performance was evaluated by the area under the receiver operating characteristic curve (AUROC), using the continuous maximum dome height as the prediction score, and by sensitivity, specificity, positive predictive value, negative predictive value, and Cohen’s *κ* at the pre-specified 50 µm cutoff. Confidence intervals for sensitivity, specificity, positive predictive value, and negative predictive value were calculated using the Wilson score method. Confidence intervals for *κ* and AUROC were estimated by eye-level bootstrap resampling with 2,000 iterations using the percentile method. The adjudicated-reference *κ* and inter-grader *κ* were compared descriptively, without a formal test of superiority, and directional disagreement between the graders was assessed by the McNemar test.

To separate detection from measurement accuracy, scan-level dome height agreement was evaluated among eyes classified as DSM-positive by both the algorithm and the corresponding grader. Within these concordant-positive eyes, B-scans were included when both methods produced a nonzero height at the same meridian. Algorithm false-negative eyes and same-meridian pairs with no valid algorithmic height were excluded from the height comparison and accounted for in the eye-level detection analysis. Agreement was reported as ICC (two-way random effects, single-measure, absolute agreement),^24^ Bland-Altman bias and 95% limits of agreement,^25^ and MAE with bootstrap 95% CIs based on 2,000 paired resamples using the percentile method. Pearson correlation and a test for proportional bias (linear regression of the difference on the mean) were also computed. Inter-grader agreement between the two graders was computed analogously on scans independently selected by both.

Morphological outputs were summarized descriptively in the original 50-eye DSM-positive subset. Clinical biometric variables were retained for cohort characterization but were not used to establish clinical associations or causal interpretations in this methods-focused analysis.

Segmentation accuracy was reported as per-scan mean absolute boundary error of the derived RPE outer border on both the OCTA-500 held-out split and the 50-scan pediatric test subset.

All statistical analyses were performed using Python (version 3.13.5) with SciPy (version 1.15.3), statsmodels (version 0.14.4), and scikit-learn (version 1.6.1). For the McNemar test and the test for proportional bias, a 2-sided *P* value < 0.05 was considered statistically significant.

## Results

### Study Cohort and Reference Standard Reliability

The final analytic cohort comprised 300 eyes from 300 pediatric patients (one eye per patient), partitioned into a 200-eye development set and a 100-eye independent test set. Table 1 summarizes baseline characteristics stratified by DSM status.

**Table 1:** Baseline characteristics of the study cohort stratified by DSM status.

| Variable | All ($n = 300$) | DSM+ ($n = 50$) | DSM- ($n = 250$) | $p$ |
| --- | --- | --- | --- | --- |
| *Demographics* |  |  |  |  |
| Age, years | $11.8 \pm 3.3$ | $10.3 \pm 3.4$ | $12.0 \pm 3.2$ | 0.004 |
| Sex, male, $n/N$ (%) | 164/299 (54.8) | 33/50 (66.0) | 131/249 (52.6) | 0.114 |
| *Refractive parameters* |  |  |  |  |
| Spherical equivalent, D | $-6.5 \pm 2.6$ | $-5.8 \pm 3.7$ | $-6.7 \pm 2.3$ | 0.012 |
| BCVA, logMAR | $0.04 \pm 0.09$ | $0.07 \pm 0.14$ | $0.03 \pm 0.08$ | 0.008 |
| *Ocular biometry* |  |  |  |  |
| Axial length, mm | $26.1 \pm 1.3$ | $25.7 \pm 1.5$ | $26.2 \pm 1.3$ | 0.049 |
| Keratometry K1, D | $42.2 \pm 1.4$ | $41.9 \pm 1.4$ | $42.3 \pm 1.4$ | 0.104 |
| Keratometry K2, D | $44.1 \pm 1.6$ | $44.3 \pm 2.1$ | $44.1 \pm 1.6$ | 0.751 |
| Anterior chamber depth, mm | $3.7 \pm 0.3$ | $3.7 \pm 0.5$ | $3.7 \pm 0.3$ | 0.210 |
| *Other* |  |  |  |  |
| Intraocular pressure, mmHg | $15.8 \pm 3.1$ | $15.3 \pm 3.0$ | $15.9 \pm 3.2$ | 0.171 |

Continuous variables are reported as mean ± SD for ease of comparison with prior pediatric biometry literature. Summary statistics and comparisons used available data. Available-case denominators (all/DSM+/DSM-) were 275/42/233 for age, axial length, and anterior chamber depth, 273/41/232 for spherical equivalent, 271/40/231 for BCVA, 261/36/225 for K1 and K2, 266/40/226 for intraocular pressure, and 299/50/249 for sex. Percentages were calculated using nonmissing values. Because the DSM-positive and DSM-negative groups were assembled by design rather than as random samples, $p$ -values are descriptive only. Continuous variables were compared using the Mann-Whitney $U$ test, which is robust to the non-normal distributions and unequal subgroup sizes in this case-enriched design. Sex was compared using the chi-square test. BCVA = best-corrected visual acuity (logMAR scale, higher values indicate worse vision). K1 = flat meridian keratometry. K2 = steep meridian keratometry.

Before evaluating the automated pipeline, we first characterized the reliability of manual DSM assessment on the 100-eye independent test set by examining the independent judgments of the two graders (Doctor A and Doctor B) prior to adjudication. Doctor A judged 28 eyes as DSM-positive, while Doctor B judged only 17 eyes as DSM-positive. The two graders agreed on 89 of 100 eyes (17 both-positive, 72 both-negative), and all 11 disagreements were in the same direction, with Doctor A positive and Doctor B negative, yielding Cohen’s *κ* of 0.690 (95% CI 0.519 to 0.841) and a statistically significant McNemar test (*P* = 0.001) indicating a systematic direction of disagreement between the two graders. The third ophthalmologist reviewed these 11 discordant eyes and adjudicated all of them as DSM-positive, consistent with Doctor A. This yielded a final reference standard of 28 DSM-positive and 72 DSM-negative eyes. Because every disagreement was resolved in the direction of Doctor A, the final reference labels were numerically identical to Doctor A’s labels.

### Automated DSM Detection and Measurement

Using the pre-specified 50 µm dome height threshold, the automated pipeline identified 23 of the 100 test-set eyes as DSM-positive. Against the adjudicated reference standard (28 DSM-positive, 72 DSM-negative), the pipeline achieved an AUROC of 0.927 (95% CI 0.850 to 0.986), sensitivity of 82.1% (64.4 to 92.1), specificity of 100% (94.9 to 100), and Cohen’s *κ* of 0.869 (0.749 to 0.972). As a sensitivity analysis against Doctor B alone (Table 2), the pipeline achieved sensitivity of 100% (81.6 to 100) at specificity of 92.8% (85.1 to 96.6). The ROC curves are shown in Figure 4.

**Figure 4:**
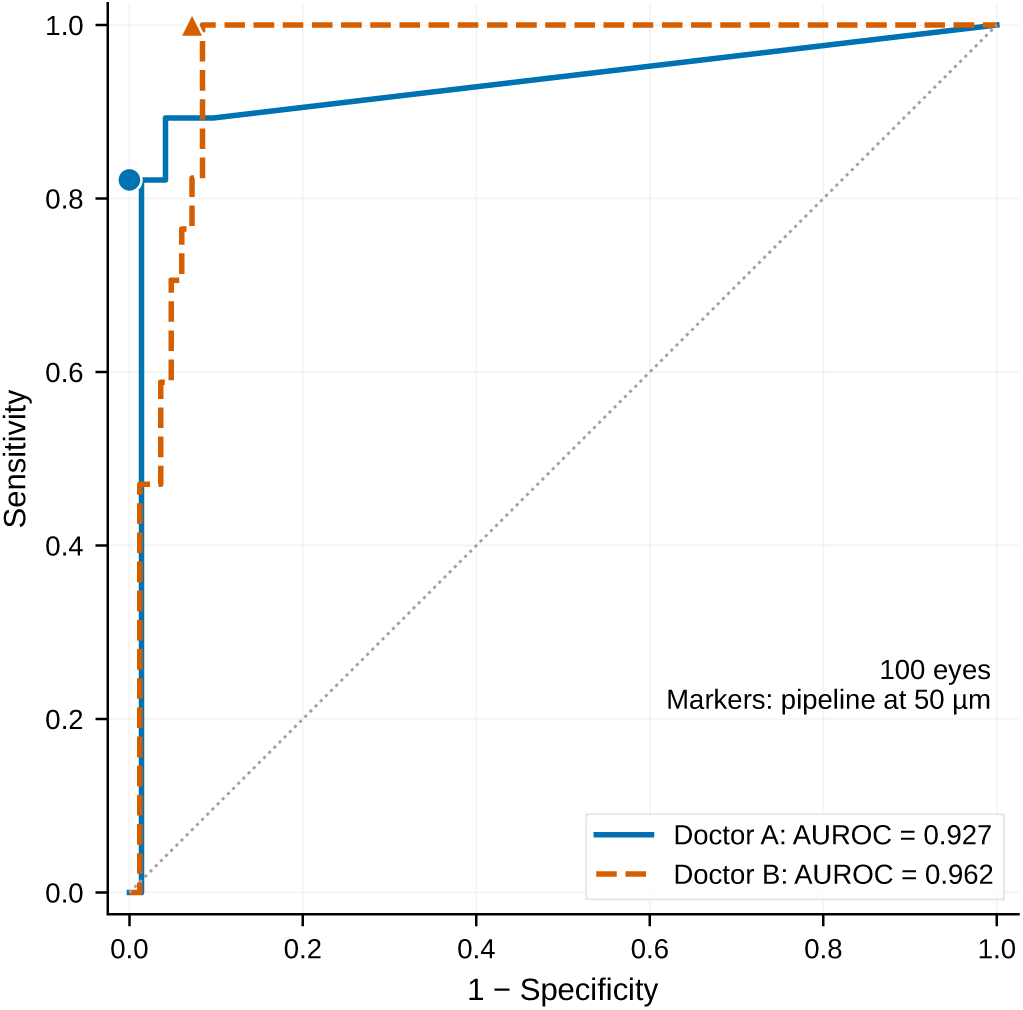
ROC curves for automated DSM detection at the eye level using the continuous maximum dome height as the prediction score. Performance is shown against Doctor A (solid blue, identical to the adjudicated reference, see Results) and Doctor B (dashed orange). Filled markers indicate the operating point at the 50 µm threshold. AUROC = area under the receiver operating characteristic curve.

**Table 2:** Automated DSM detection performance (eye-level, *n* = 100). The algorithm’s operating threshold was 50 µm.

| Metric | Adjudicated reference* | Doctor A | Doctor B |
| --- | --- | --- | --- |
| TP / FP / FN / TN | 23 / 0 / 5 / 72 | 23 / 0 / 5 / 72 | 17 / 6 / 0 / 77 |
| Sensitivity (95% CI) | 82.1% (64.4 to 92.1) | 82.1% (64.4 to 92.1) | 100% (81.6 to 100) |
| Specificity (95% CI) | 100% (94.9 to 100)<sup>†</sup> | 100% (94.9 to 100) | 92.8% (85.1 to 96.6) |
| PPV (95% CI) | 100% (85.7 to 100) | 100% (85.7 to 100) | 73.9% (53.5 to 87.5) |
| NPV (95% CI) | 93.5% (85.7 to 97.2) | 93.5% (85.7 to 97.2) | 100% (95.2 to 100) |
| Cohen’s $\kappa$ (95% CI) | 0.869 (0.749 to 0.972) | 0.869 (0.749 to 0.972) | 0.814 (0.652 to 0.940) |
| AUROC (95% CI) | 0.927 (0.850 to 0.986) | 0.927 (0.850 to 0.986) | 0.962 (0.922 to 0.991) |

TP = true positive. FP = false positive. FN = false negative. TN = true negative. PPV = positive predictive value. NPV = negative predictive value. $\kappa$ = Cohen’s kappa. AUROC was computed using the continuous maximum dome height ( $\mu\text{m}$ ) as the prediction score. Ninety-five percent confidence intervals for sensitivity, specificity, PPV, and NPV were computed using the Wilson score method. CIs for $\kappa$ and AUROC were estimated by eye-level bootstrap resampling with 2,000 iterations using the percentile method. \*The adjudicated reference was the primary reference standard; it was numerically identical to Doctor A’s labels because all 11 inter-grader disagreements were adjudicated in the direction of Doctor A, so the first two columns are identical. Results against Doctor B are shown as a sensitivity analysis. <sup>†</sup>The 100% specificity versus the adjudicated reference reflects the algorithm’s conservative operating point because all 23 algorithm-positive eyes were also labeled positive by Doctor A, whereas Doctor A identified more DSM-positive eyes than the algorithm.

The algorithm produced no false positives against the adjudicated reference standard. All 23 eyes that the algorithm flagged as DSM-positive were also DSM-positive in the reference standard. The algorithm missed 5 eyes that the reference standard classified as DSM-positive, and all 5 were among the 11 eyes on which Doctor A and Doctor B had originally disagreed (Doctor A positive, Doctor B negative) and that were subsequently adjudicated as DSM-positive. Of those 11 disagreement eyes, the algorithm correctly identified 6 and missed 5.

Case review identified more than one source of false-negative classification. In 1 eye with a manual height of 117.3 µm, extreme discontinuity of the segmented boundary contour triggered quality rejection before tangent-line measurement. In 3 eyes with manual heights of 55.4, 78.6, and 105.6 µm, the geometric module did not establish a valid tangent-line baseline on the grader-selected scan and therefore produced no valid same-meridian height. In the remaining eye, both methods produced valid measurements on the same scan, but the algorithmic height was below the diagnostic threshold (36.1 vs 64.4 µm manually). Representative inter-grader discordant eyes, with the corresponding algorithm outputs, are shown in Supplementary Figure S1.

To separate measurement accuracy from detection performance, height agreement was evaluated on same-meridian paired scans from eyes classified as DSM-positive by both the algorithm and the corresponding grader. Pairs were included when both methods produced a nonzero height on that scan. Detection false negatives and scans with no valid algorithmic height were accounted for in the eye-level analysis (Table 2) rather than being included as measurement errors. Because graders independently selected which scans to annotate, the number of paired measurements differed by grader. The results are summarized in Table 3, Figure 5, and Figure 6.

**Figure 5:**
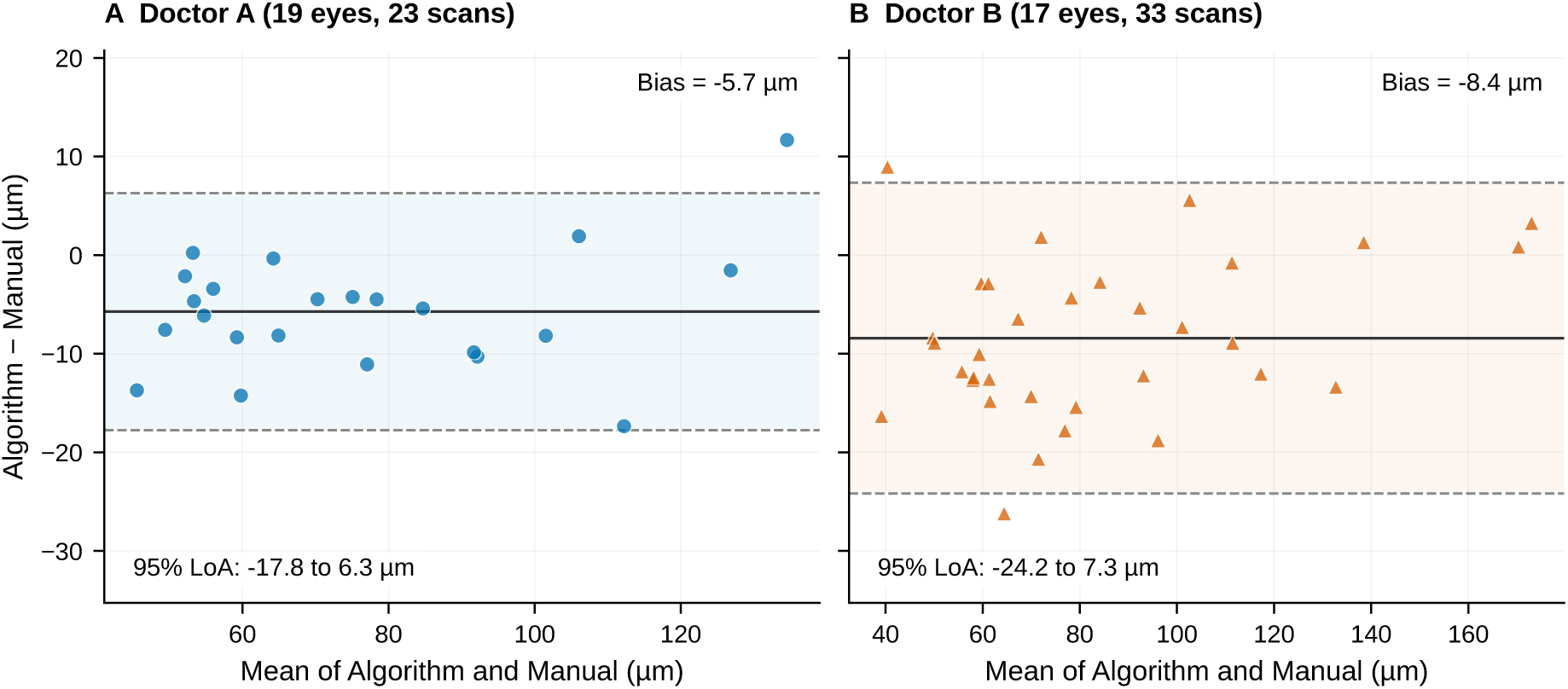
Bland-Altman plots of dome height measurements on same-meridian paired scans from concordant-positive eyes. **Left,** algorithm vs Doctor A. **Right,** algorithm vs Doctor B. Solid line, mean bias. Dashed lines and shaded band, 95% limits of agreement.

**Figure 6:**
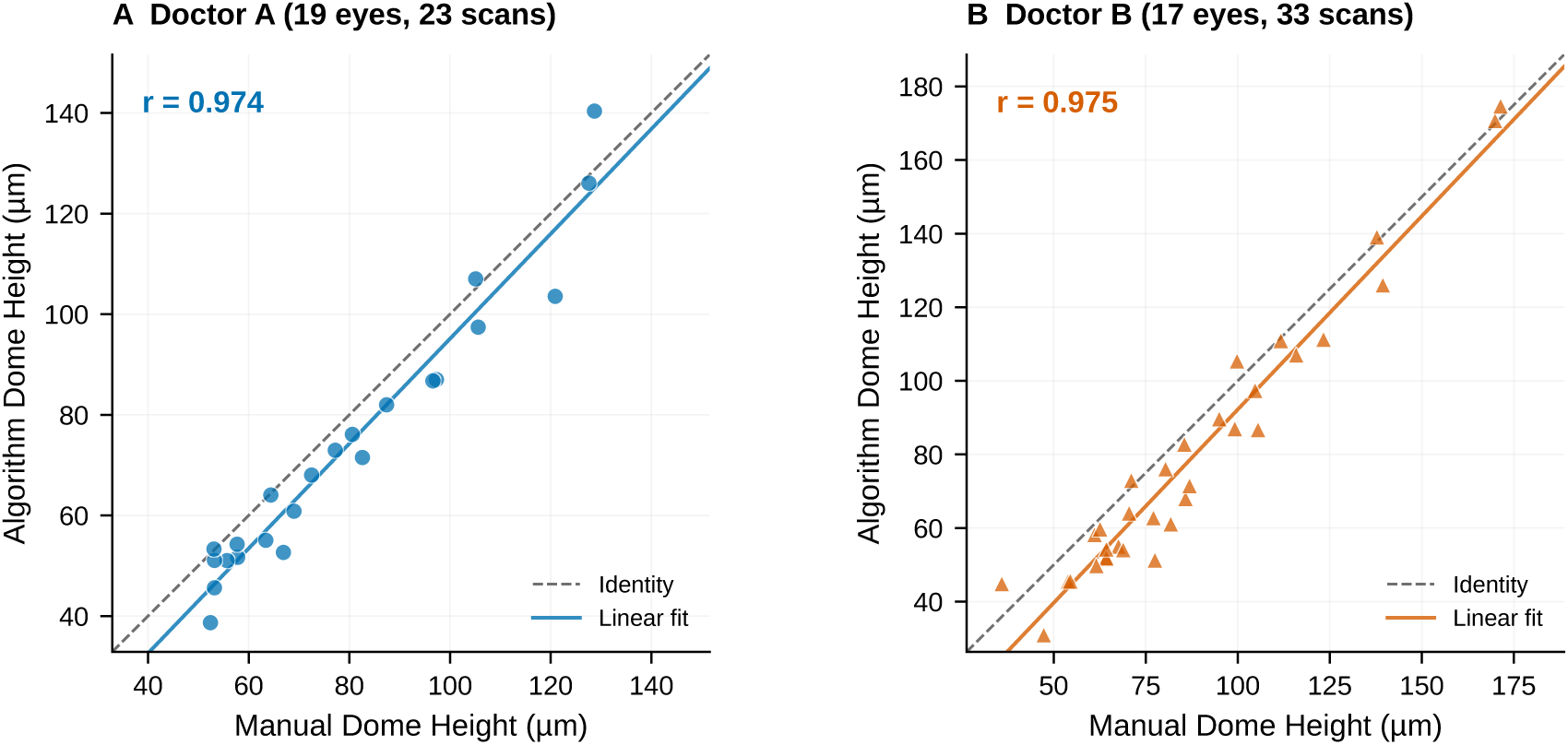
Automated vs manual dome height on same-meridian paired scans from concordantpositive eyes. **Left,** algorithm vs Doctor A. **Right,** algorithm vs Doctor B. Dashed line, identity. Solid colored line, linear regression fit.

**Table 3:** Agreement of dome height measurements on same-meridian paired scans from concordant-positive eyes. Eyes were included when both the algorithm and the corresponding grader classified them as DSM-positive, and scans were included when both methods produced a nonzero height.

| Comparison | $N$ | ICC (95% CI) | MAE ($\mu\text{m}$) | Bias ($\mu\text{m}$) (95% CI) | 95% LoA ($\mu\text{m}$) |
| --- | --- | --- | --- | --- | --- |
| Doctor A vs Doctor B | 14 | 0.953 (0.883 to 0.973) | 5.9 (3.8 to 8.1) | -4.9 (-7.7 to -2.1) | -15.5 to 5.7 |
| Algorithm vs Doctor A | 23 | 0.949 (0.899 to 0.972) | 6.9 (5.1 to 8.8) | -5.7 (-8.2 to -3.2) | -17.8 to 6.3 |
| Algorithm vs Doctor B | 33 | 0.944 (0.874 to 0.971) | 9.7 (7.6 to 11.9) | -8.4 (-11.2 to -5.7) | -24.2 to 7.3 |

ICC = intraclass correlation coefficient (two-way random, single-measure, absolute agreement, ICC(2,1) in Shrout-Fleiss notation). MAE = mean absolute error (95% CI by paired bootstrap, 2,000 iterations). Bias = algorithm - manual, with 95% CI computed as $\bar{d} \pm 1.96 \cdot \text{SD}_d / \sqrt{N}$ . LoA = limits of agreement. $N$ = number of same-meridian paired scans from eyes classified as DSM-positive by both the algorithm and the corresponding grader, with a nonzero height produced by both methods. Algorithm values use the physical-coordinate height calculation described above. Algorithm false-negative eyes and pairs with no valid algorithmic height were excluded from this table and accounted for in the detection performance analysis (Table 2).

The automated measurements showed a small negative bias relative to manual measurements: *−*5.7 µm versus Doctor A and *−*8.4 µm versus Doctor B (Figure 5). These biases correspond to roughly 2 to 3 axial sampling intervals at the DRI OCT Triton’s axial sampling of 2.61 µm per pixel. Pearson correlations between automated and manual measurements remained high (0.974 against Doctor A and 0.975 against Doctor B, Figure 6); correlation was interpreted as a descriptive complement to agreement rather than as evidence of equivalence.

### Morphological Quantification and Supporting Segmentation Accuracy

The automated pipeline extracted the three core morphological features for all 50 DSM-positive eyes in the study cohort. Using the physical-coordinate height calculation, maximum dome height was 87.7 *±* 40.8 µm (median 74.4, IQR 57.9 to 105.1), maximum projected base width was 5404 *±* 1322 µm (median 5660, IQR 4986 to 6102), and the steepness index was 0.017 *±* 0.009 (median 0.014, IQR 0.012 to 0.020).

Among the 50 DSM-positive eyes, 34 (68%) showed a horizontal pattern, 5 (10%) vertical, 4 (8%) bidirectional, and 7 (14%) oblique. Only 1 eye (2%) reached 50 µm on all 12 meridians, meeting the complete-dome definition of García-Zamora et al.;^9^ in the remaining eyes, the threshold-crossing elevation was confined to a subset of meridians, consistent with ridge-shaped configurations. Representative patterns are shown in Figure 2; no inferential comparison between orientation subgroups was performed.

All of the above detection and measurement results depend on the accuracy of the underlying RPE outer border delineation. We therefore evaluated the segmentation model’s RPE outer border accuracy on two datasets. On the OCTA-500 held-out split of 24,000 B-scans, the RPE outer border MAE was 0.69 *±* 1.03 µm. On the 50-B-scan pediatric test subset drawn from the 100-eye independent test set, the RPE outer border MAE was 1.54 *±* 0.87 µm (Table 4).

**Table 4:**
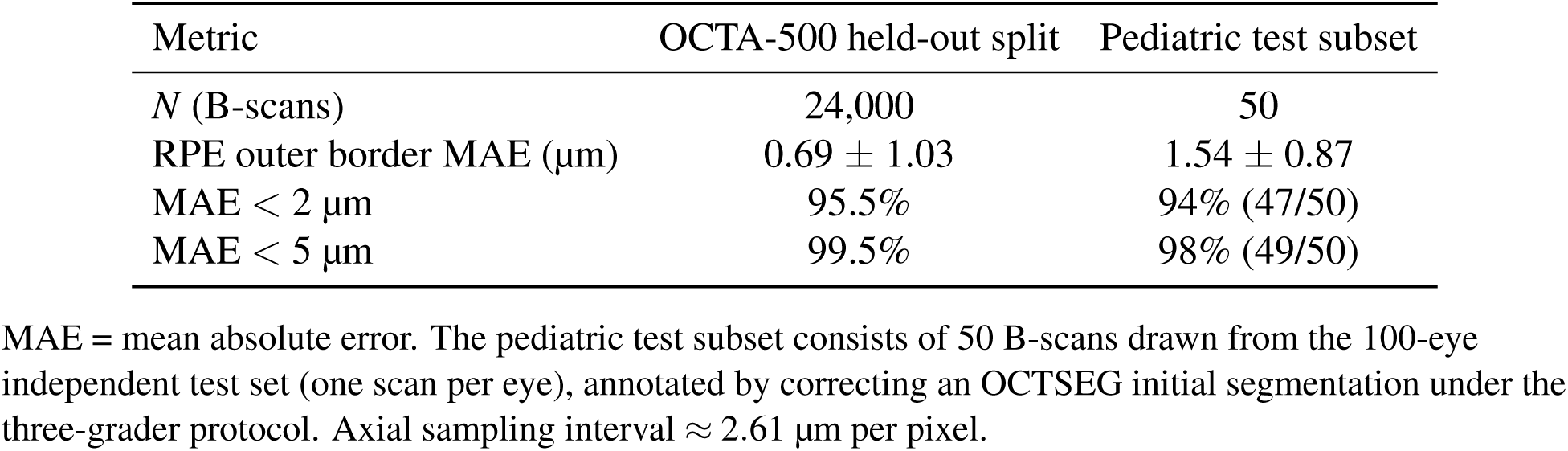
RPE outer border segmentation accuracy on the OCTA-500 held-out split and on the pediatric test subset drawn from the 100-eye independent test set.

| Metric | OCTA-500 held-out split | Pediatric test subset |
| --- | --- | --- |
| $N$ (B-scans) | 24,000 | 50 |
| RPE outer border MAE ($\mu\text{m}$) | $0.69 \pm 1.03$ | $1.54 \pm 0.87$ |
| MAE $< 2 \mu\text{m}$ | 95.5% | 94% (47/50) |
| MAE $< 5 \mu\text{m}$ | 99.5% | 98% (49/50) |

MAE = mean absolute error. The pediatric test subset consists of 50 B-scans drawn from the 100-eye independent test set (one scan per eye), annotated by correcting an OCTSEG initial segmentation under the three-grader protocol. Axial sampling interval $\approx 2.61 \mu\text{m}$ per pixel.

## Discussion

In this study, we developed an interpretable automated pipeline that detects and measures DSM from OCT B-scans and evaluated it against masked expert grading in a pediatric high-myopia test bed. The pipeline reproduced the adjudicated eye-level classifications with high specificity, and its height measurements agreed with each grader to within a few axial sampling intervals. Because the adjudicated reference labels were identical to Doctor A’s labels, the higher algorithm-versus-reference *κ* relative to inter-grader *κ* is descriptive and does not establish algorithmic superiority.

These findings address a recognized challenge in DSM assessment. Two experienced graders in our cohort reached different binary judgments in 11 of 100 eyes, and every disagreement was in the same direction, with Doctor A positive and Doctor B negative. This pattern suggests different diagnostic operating thresholds between graders. The adjudicated reference represents expert consensus rather than an independent gold standard, because the third grader reviewed only discordant eyes and all adjudications agreed with Doctor A. A deterministic, interpretable automated tool can serve as a complement to clinical judgment rather than a replacement, an approach similar to recent automated OCT quantification pipelines for the anterior segment,^13^ choroid,^14^ and geographic atrophy.^12^ In a potential triage model, the algorithm could handle clearly positive and clearly negative cases, allowing clinician review to focus on ambiguous cases; this workflow remains to be evaluated prospectively. With processing times on the order of milliseconds per B-scan on a single consumer-grade GPU, speed is unlikely to limit use during routine outpatient visits. In large-scale school screening such as SCALE-HM, the interpretable report (segmented RPE outer border, tangent-line baseline, perpendicular height) may allow readers to verify or flag automated findings efficiently, although its use by non-specialist readers was not tested in this study.

Two design choices distinguish our pipeline from prior automated DSM work. First, it exploits the full 12-meridian radial OCT acquisition. Existing methods such as the DSMC index of Del Fabbro et al.^10^ still require manual landmark identification and a model-based geometric summary, while end-to-end deep learning classifiers such as that of Ye et al.^11^ output only a binary label with no anatomical traceback. Evaluating all 12 meridians allows our pipeline to identify the maximum dome height across orientations, record a meridional orientation pattern, and output a richer structural description (maximum dome height, projected base width, and steepness index) rather than a single scalar detection score. Second, every measurement traces back to identifiable landmarks (segmentation, baseline, contact points, apex) on the source B-scan, so the same input yields the same numeric output and clinicians can verify each measurement against the source anatomy. The interpretability offered here is therefore geometric and anatomical: it recovers quantities that can be checked on the source image, rather than merely highlighting salient image regions. Table 5 compares the present approach with previous DSM assessment methods.

**Table 5:** Comparison of the present approach with previous DSM assessment methods.

| Study | Population (*n*) | OCT protocol | DSM detection | Height measurement | Reported agreement or performance |
| --- | --- | --- | --- | --- | --- |
| Caillaux et al.<sup>6</sup> | Adult myopic eyes with DSM (48 eyes) | Horizontal and vertical SD-OCT, 3D map | Manual | Manual caliper | – |
| Shin et al.<sup>17</sup> | Children and adolescents (1,042 eyes; 8 with DSM) | Horizontal and vertical SD-OCT | Manual | Manual caliper | – |
| García-Zamora et al.<sup>9</sup> | Highly myopic eyes (49 eyes) | 12-line radial SS-OCT | Manual | Manual | – |
| Dang et al.<sup>26</sup> | Preterm and full-term infants (217 sessions, 80 infants) | Handheld SS-OCT | Manual grading | Semi-automated (grader-selected points) | – |
| Del Fabbro et al.<sup>10</sup> | Myopic eyes with DSM (125 eyes) | SD-OCT | Manual | Manual landmarks, DSMC index | Inter-grader ICC 0.96 (height) |
| Ye et al.<sup>11</sup> | High myopia (450 test images) | Single B-scan images | Automated (end-to-end CNN) | Not provided | AUC 0.955 (DSM) |
| Present study | Children with high myopia (100 test eyes) | 12-line radial SS-OCT | Automated | Automated, traceable to landmarks | AUROC 0.927; ICC 0.949 vs grader |

Metrics are shown as reported and are not directly comparable across studies because of differences in population, device, reference standard, and outcome definition. SD-OCT = spectral-domain OCT. SS-OCT = swept-source OCT. DSMC = dome-shaped macula curvature. CNN = convolutional neural network. AUC = area under the curve. ICC = intraclass correlation coefficient.

The pipeline is also fully deterministic. Because it operates without test-time augmentation, ensemble averaging, or any other stochastic component, repeated processing of the same B-scan returns the same dome height to machine precision. The bias and limits of agreement reported above against expert graders therefore reflect inter-method differences, not run-to-run model variation. This property supports reproducible reanalysis of a fixed image; repeat-scan studies are still needed to quantify clinical measurement repeatability across image acquisition.

The false-negative analysis also indicates when automated output should be reviewed. Four of the 5 false negatives occurred in scans that the pipeline itself flagged, either through contour-quality rejection or through failure to establish a valid tangent-line baseline. Because these states are reported explicitly for each meridian, a clinical workflow could accept clearly positive and clearly negative outputs while routing eyes with rejected or unmeasurable meridians, and eyes with maximum heights near the 50 µm cutoff, to manual review. The review criteria and their effect on workload would need to be defined prospectively.

The 50-eye morphology subset was used to describe the range of outputs rather than to establish disease mechanisms or clinical biomarkers. The mean maximum dome height was 87.7 µm, below the 407.7 µm reported by Caillaux et al.^6^ in a referral series of adult myopic eyes with DSM; this difference is descriptive and may reflect population, sampling, and measurement-definition differences. The predominance of horizontal patterns (68%) is consistent with previous reports in adults and children.^6,27^ The pipeline characterizes the RPE protrusion but does not image the scleral features needed to distinguish proposed mechanisms of DSM.^5,28^

This study has several limitations. First, it is a single-center retrospective analysis conducted on a single OCT platform (DRI OCT Triton, swept-source). The U-Net backbone was pre-trained on the external OCTA-500 dataset and fine-tuned on internal pediatric annotations, which provides one layer of cross-domain transfer. However, the end-to-end DSM detection-and-measurement pipeline itself has only been evaluated on a held-out internal subset of the same SCALE-HM cohort. External validation across additional institutions, OCT devices (spectral-domain and other swept-source platforms), age groups, and acquisition protocols will therefore be needed before broader clinical deployment. Second, the 50 µm dome-height threshold is widely used in adult and pediatric OCT studies but is not universally standardized.^5,17^ The 5 false negatives did not represent a uniform group of threshold-adjacent measurements. They included 1 contour-quality rejection before tangent-line measurement, 3 cases in which the geometric module could not establish a valid tangent-line baseline on the grader-selected scan, and 1 valid algorithmic measurement below the threshold. Third, the adjudicated reference was structurally dependent on Doctor A because all 11 inter-grader disagreements were resolved in the direction of Doctor A, making the final reference labels numerically identical to that grader’s labels. The third grader reviewed only discordant eyes, so the design could not detect errors shared by the 2 initial graders in concordant cases. Algorithm-vs-reference and inter-grader agreement estimates should therefore be interpreted descriptively rather than as a direct comparison of algorithmic and human diagnostic ability. Fourth, we used DSM as an umbrella OCT phenotype. Under a stricter 12-line radial definition, configurations confined to selected meridians may be classified as RSM; the recorded orientation patterns should therefore not be interpreted as proof of complete round three-dimensional dome morphology.^9^ Fifth, the morphology analysis was a descriptive internal analysis of a 50-eye case-enriched subset and was not designed to estimate population prevalence, test clinical associations, or establish longitudinal change. Sixth, a single set of geometric rules for apex and baseline construction was evaluated; alternative geometric definitions and repeat-scan reliability remain to be assessed.

Several extensions follow from this work. Evaluation on other scan patterns, including dense raster volumes, would allow three-dimensional reconstruction of dome morphology. Application to longitudinal acquisitions could quantify dome height change during childhood myopia progression. External evaluation across devices, centers, and age groups, including adults, is needed to establish generalizability.

## Conclusions

Together, these results show that an interpretable automated pipeline based on radial OCT can detect DSM and quantify dome morphology, with internal agreement against the adjudicated expert reference and both individual graders demonstrated in a pediatric high-myopia test bed. External, cross-device, cross-age, and longitudinal evaluation remains necessary before clinical deployment.

## Data Availability

Model produced are available online at: https://github.com/MaybeRichard/Automatic-Dome-shaped-Macula-Detection

## Acknowledgments

Supported by the Basic Research Program of Shanghai Eye Disease Prevention and Treatment Center (Grant No. 2021JC002), the Clinical Research Program of Shanghai Eye Disease Prevention and Treatment Center (Grant No. 22LC01009), and the Tongji University Medicine + X Interdisciplinary Research Program (Grant No. 2026-YB-0255C-01).

Disclosure: F. Yang, None; Y. Zhou, None; B. Zhang, None; J. Yang, None; L. Du, None; C. Yu, None; X. Chen, None; J. Zhu, None; Y. Wang, None; X. He, None.

## Data and Code Availability

Source code for the segmentation U-Net (training and inference), the geometric DSM detection pipeline, and the evaluation and analysis scripts used in this study is publicly available at https://github.com/MaybeRichard/Automatic-Dome-shaped-Macula-Detection under the MIT license. The OCTA-500 dataset used for segmentation pre-training is publicly available from its original source.^21^ The pediatric SCALE-HM cohort analyzed in this study is governed by institutional data-sharing agreements and is not publicly redistributable. De-identified summary data are available from the corresponding author upon reasonable request, subject to institutional review board approval.

